# Responding to disruption: Exploring the transition to telehealth in mental-health occupational therapy during the COVID-19 pandemic

**DOI:** 10.1101/2022.09.19.22280127

**Authors:** Aislinn Duffy, Bryan Boyle, Eoin Gorman, Sarah Hayes

## Abstract

**Background:** COVID-19 presented significant challenges for occupational therapy (OT) services in Ireland. Public health guidelines necessitated a transition of services from face-to-face delivery to the use of telehealth modalities. Telehealth has yet to be extensively researched within mental health OT, with a particular need for an increased understanding of therapeutic processes when conducted remotely.

**Aim:** To explore the experiences of occupational therapists transitioning to telehealth service provision.

**Material and Methods:** This study employed a qualitative, descriptive design to examine the experiences of therapists transitioning from face-to-face to telehealth services within a mental health service. Data was collected using comprehensive, semi-structured interviews with four participants and analysed thematically.

**Results:** This study yielded three major themes: 1) responding to disruption, 2) reconsidering practice with technology and 3) therapeutic use of the ‘virtual self’.

**Conclusions:** Adaptation to telehealth provision requires planned, gradual transition but offers unique opportunities for therapeutic engagement. How space is considered in therapy as well as therapists’ communication styles are components of practice which are altered when conducted remotely.

**Significance:** The disruption caused by COVID-19 presented opportunities for considering the delivery of OT services. As services emerge from social restrictions it is likely that their recent experiences will be utilised in reconfiguring the future delivery of mental-health OT services.

## Introduction

Occupational therapists have a distinct role in promoting the mental health and wellbeing of individuals and populations. The World Federation of Occupational Therapists [WFOT] (2019) states that occupational therapists may work to promote well-being in their clients, prevent the development of mental illnesses and promote recovery in clients who have experienced mental health difficulties. Occupational therapists deliver interventions aimed at improving clients’ mental wellbeing in community mental health settings (American Occupational Therapy Association [AOTA], 2016). Interventions are delivered on an individual, group, or population level, and may involve education, self-management, and the promotion of engagement in valued occupations (Cole, 2014; McCullough, 2014; WFOT, 2019).

As a result of the COVID-19 pandemic, services provided by many occupational therapists in Ireland were disrupted due to restrictive public health guidelines, with many clients receiving reduced levels of service provision (Association of Occupational Therapists of Ireland [AOTI], n.d.), including those in community mental health settings. Telehealth has been suggested to provide an alternative to in-person service delivery when face-to-face sessions cannot be facilitated (WFOT, 2019). Telehealth refers to *‘the application of evaluative, consultative, preventative, and therapeutic services delivered through information and communication technology’* (AOTA, 2018, p.1). Considering the disruptions to services resulting from the COVID-19 pandemic, telehealth provided therapists with an opportunity to continue to deliver services to their clients in a safe manner (Sarsak, 2020). Recent research indicates that the transition to telehealth across occupational therapy practices will have lasting effects on the profession worldwide, with increased technology use persisting as a component of practice going forward (Hoel et al., 2021).

As early as 2000, telehealth was identified as an important next step within occupational therapy, as outlined in the Canadian Association of Occupational Therapists’ position statement on telehealth (2000). Telehealth has subsequently been implemented across a variety of settings within occupational therapy practice, including physical rehabilitation (Harkey et al., 2020; Tenforde et al., 2017) and paediatrics (Zylstra, 2013), as well as across various disciplines within healthcare, such as physiotherapy (Wong et al., 2020) and nursing (Taylor et al., 2014). The level of complexity of the technology used in telehealth implementation within occupational therapy ranges from simple, commonly used modalities such as phone calls (Dahl-Popolizio, 2020), to rarer and more complex modalities, such as 3D simulations of clients’ homes to support the planning of home modifications (Atwal et al., 2013). Furthermore, the Department of Health’s (2019) strategy for eHealth indicates the rising pertinence of information and communications technology development within healthcare services in Ireland. The implementation of telehealth within services requires a process of transition, during which services acquire the resources and skills required for the delivery of remote therapy. Factors such as transition costs and negative perceptions of technology use in practice may pose as barriers to this transition (Tsai et al., 2019).

The purpose of this research study was to explore the experiences of occupational therapists in their transition to telehealth use in a community mental health setting. A qualitative, descriptive approach was used to best capture participants’ experiences of this transition. As telehealth in Ireland is still in the early stages of development (Lolich et al., 2019), investigation is needed to explore therapists’ experiences of telehealth across occupational therapy settings to inform future practice in this expanding area.

### Literature Review

Searches of EBSCOHost, PubMed and Scopus databases were completed to identify literature related to the use of telehealth in occupational therapy practice. The key search terms used included ‘occupational therapy OR occupational therapist OR OT’ as well as ‘telehealth OR telemedicine OR telemonitoring OR telepractice OR telecare OR ehealth’. Original searches included terms relating to mental health, but this was then excluded as results were narrowed greatly. Articles containing these key terms in their title or abstract were identified. All searches were limited to peer-reviewed, full text articles, published within the last ten years. The researchers examined the abstracts of the 156 articles yielded through the database searches. The reference lists of relevant articles were also examined to identify further relevant literature which did not appear in the original database search. Twenty-five articles were identified which related specifically to telehealth in occupational therapy practice and were included in the literature review. Recurring themes within the literature included the increased efficiency and accessibility of services due to telehealth implementation, technology associated difficulties, and the impact of telehealth on the therapeutic relationship.

### Increased Efficiency of Services

The potential for the implementation of telehealth to improve the efficiency of paediatric and adult physical services was identified within the literature. No literature was identified relating to mental health services. Within individual physical services, clinicians acknowledged the potential for telehealth to address clients’ needs which are currently not being met by existing services within primary care and outpatient clinics (Lolich et al., 2019; Morris et al., 2019), such as the provision of additional support between on-site visits.

Telehealth has also been recognised to offer opportunities for increased efficiency in the sharing of information between professionals and the management of administrative tasks within primary care services (Lolich et al., 2019). Furthermore, telehealth reduces the amount of travel required by therapists and clients during service provision in discharge planning from acute settings and school-based occupational therapy practice, which has subsequent implications for cost reduction and increased time efficiency (Atwal et al., 2013; Rortvedt & Jacobs, 2019). It has been suggested that due to this increased efficiency, the implementation of telehealth may prevent delays in service provision for clients (Atwal et al., 2013; Cason, 2014).

### Increased Accessibility of Services

Within the literature, both practitioners and service users reported increased accessibility of services through use of telehealth within adult and paediatric practice in the community (Hines et al., 2019; Renda & Lape, 2018). Delivery of services via telehealth increased accessibility for service users living in rural and underserved areas of Australia, the United States and Denmark, due to reduced travel demands (Hines et al., 2019; Kuravackel et al., 2018; Little et al., 2018; Rosenbek Minet et al., 2015; Scriven et al., 2019). Telehealth has also been shown to increase flexibility in the scheduling of sessions within primary care settings where home visits are routinely required to deliver advice on home modifications (Renda & Lape, 2018). Telehealth also increases flexibility in the length and location of sessions offered to clients within paediatric services, facilitating increased levels of client-centredness in practice (Hines et al., 2019). Improved accessibility supports equity in healthcare, as those experiencing chronic conditions who cannot access transport may attend rehabilitation services remotely (Rosenbek Minet et al., 2015). Overall satisfaction with telehealth services (Hines et al., 2019; Iacono et al., 2016), and positive therapy outcomes using telehealth (Dunleavy et al., 2013; Iacono et al., 2016; Kuravackel et al., 2018; Little et al., 2018; Nissen & Serwe, 2018) have been reported by both practitioners and service users within physical community services for both adults and children in individual and group settings.

### Technology Associated Difficulties

Despite the purported benefits, numerous challenges associated with telehealth implementation have been identified. Frequent technical difficulties with telehealth infrastructure experienced by both healthcare professionals and service users have been reported internationally within the literature (Breeden, 2016; Dunleavy et al., 2013; Gately et al., 2020; Reifenberg et al., 2017; Renda & Lape, 2018; Rortvedt & Jacobs, 2019; Stillerova et al., 2016). Research suggests access to the required technology and to broadband of sufficient quality can pose as barriers to telehealth implementation (Breeden, 2016; Hines et al., 2019; Morris et al., 2019). Additionally, concerns have been identified about choosing a software that protects the security and privacy of clients, which is an additional challenge in utilising telehealth technology in school-based occupational therapy practice (Rortvedt & Jacobs, 2019). Concerns for privacy and data protection have posed as a deterrent to telehealth use in the United States and in Ireland (Cason et al., 2012; Lolich et al., 2019; Rortvedt & Jacobs, 2019), and this was noted as one of the primary causes of delay in the uptake of telehealth in Ireland (Lolich et al., 2019). Given these concerns, ensuring the use of secure and private platforms to deliver therapy may facilitate the uptake of telehealth and ease the apprehensions of therapists and service users (Cason et al., 2012).

Proficiency of technology use has presented difficulties for both occupational therapists and service users, with therapists expressing concerns that they have insufficient knowledge in relation to telehealth, as well as concerns that clients would have difficulty with telehealth technology use (Morris et al., 2019). While service users reported comfort in technology has been noted within the literature, including older adults in community settings and parents of children with ASD in paediatric practice (Breeden, 2016; Ingersoll et al., 2017), it was estimated in 2017 that 46% of Irish people between the ages of 60 and 74 had no previous experience of internet use, which may pose challenges in utilising telehealth with this population (Lolich et al., 2019). Guise and Wiig (2017) outlined the need for appropriate training to combat negative attitudes of professionals towards telehealth implementation, as well as to increase their knowledge in delivering therapy effectively in this way. Therapist apprehension and a perceived need for increased telehealth training have been shown to act as challenges to successful telehealth implementation (Atwal et al., 2013; Iacono et al., 2016; Lolich et al., 2019; Morris et al., 2019; Rortvedt & Jacobs, 2019).

### Impact on Therapeutic Relationship

The feasibility of the development and maintenance of therapeutic relationships using telehealth is a common theme within the literature. Within the literature the therapeutic use of self is described as a complex process in which occupational therapists use *‘professional reasoning, empathy, and a client-centered, collaborative approach’* (p. 20) to develop and maintain therapeutic relationships with service users (AOTA, 2020). The development of rapport and a strong therapeutic relationship, as well as building trust with clients and their families have been identified as important factors in the successful implementation of telehealth within occupational therapy practice in community settings (Breeden, 2016; Gardner et al., 2016). However, it has been reported that professionals find it difficult to develop rapport with service users when delivering therapy via telehealth, and that the building of rapport in this way requires intentional effort and skill (Breeden, 2016; Rortvedt & Jacobs, 2019). Rortvedt and Jacobs (2019) presented the views of professionals working in a school setting in the U.S., including OTs, district administrators and technology staff, who expressed concerns regarding the viability of the therapeutic relationship in the context of telehealth due to the absence of *“human connection”* (p. 129). Technology related factors such as poor broadband connection, poor audio and video quality, and the need to address these technical issues disrupt sessions and negatively impact upon session quality (Breeden, 2016; Dunleavy et al., 2013; Reifenberg et al., 2017; Renda & Lape, 2018; Stillerova et al., 2016), which has negative implications for the therapeutic relationship and the development of therapeutic rapport. Within Garner et al’s (2016) study, parents of children with developmental disabilities highlighted varied opinions on the ability to develop therapeutic relationships with occupational therapists remotely. Within this study, some participants stated that they felt they could develop therapeutic relationships with OTs remotely, while others stated their preference for first developing the therapeutic relationship with the OT in-person before utilising remote services (Gardner et al., 2016).

### Limitations in Current Research

Telehealth is an emerging area of practice within occupational therapy, with the majority of existing research published in the last five years, leaving several gaps in the literature yet to be addressed. Over half of the research identified within the literature review was conducted within the U.S., and the majority of studies related to the use of telehealth in adult and paediatric community services. No evidence was identified relating to telehealth in occupational therapy mental health settings. Hwang et al.’s systematic review (2020) demonstrated that psychosocial interventions, such as those focused on lifestyle behaviour changes and problem-solving skills, have the potential to improve symptoms of depression, fatigue, and distress, while also improving quality of life when delivered via telehealth. Despite this, no articles detailing the use of telehealth in mental health occupational therapy practice relating to the perspectives of either occupational therapists or service users were yielded in the search outlined above. Though the potential for telehealth use in mental health occupational therapy practice has been acknowledged (Cason, 2012), this area represents a gap within the current literature. Limited implementation of telehealth within mental health services in Ireland has been seen to-date, which may be due to the lack of research in this area conducted in an Irish context (Cullen, 2018). Literature across healthcare disciplines demonstrated that a process of transition is required to integrate telehealth into existing services (Fischer et al., 2021; Fryer et al., 2020; Nicholas et al., 2021). This process of transition involves the trial of telehealth modalities, development of new protocols, and the establishment of best practice for the service using telehealth (Fryer et al., 2020; Nicholas et al., 2021). However, this transition has yet to be explored within occupational therapy practice.

Consequently, additional research exploring the process of transition to telehealth within mental health occupational therapy is required to inform practice in this area going forward. Based on the gaps in current literature, the primary aim of this research project was to explore the experiences of occupational therapists in transitioning to telehealth during the COVID-19 pandemic within community mental health services. Additional aims included to explore how the process of transition to telehealth occurs within community mental health services, identify what opportunities and challenges are presented by the integration of telehealth into a mental health service, and examine what ways occupational therapists transfer and apply their professional skills in order to successfully deliver practice through telehealth.

## Methodology Materials and Methods

A qualitative research design was implemented within this study. Qualitative research provides answers to questions that explore the experiences of the participant and is beneficial when investigating topics where the participant’s point of view has not yet been thoroughly researched (Carpenter & Suto, 2008). Given the gaps evident in the literature relating to occupational therapists’ experiences of telehealth in mental health settings, a qualitative design was most pertinent in addressing the identified research questions. This study drew from the principles of a descriptive exploratory approach in sampling, data generation and using low-inference descriptions in presenting the data, which enables validity of description and provides detailed accounts of experiences being studied (Sandelowski, 2000). This approach enabled the examination of the process of transition to telehealth, as well as the impact of this transition upon a service within a community mental health setting.

## Method

Semi-structured interviews were conducted to explore the experiences of four occupational therapists in delivering occupational therapy via telehealth in a community mental health setting. A semi-structured interview method supports the generation of detailed data relating to the participants’ perspectives (Lysack et al., 2017). Interviews are a frequently used method of data generation when using a descriptive exploratory approach (Sandelowski, 2000), as they can elicit rich information relating to participant perspectives (Polgar & Thomas, 2013), which was deemed pertinent given the lack of exploration conducted to date regarding the perspectives of Irish occupational therapists in relation to telehealth.

A set of seven questions were devised by the researchers for use within the interviews, based on the research aim and questions as well as the gaps in current literature. The questions were divided into three sections to explore 1) participants’ experience with telehealth to date, 2) opportunities and challenges presented by telehealth, and 3) participants’ experiences of the transition to telehealth. Additional probing questions were used by the researchers, based on the participants’ responses, to elicit further information. An inductive approach ensured topics generated by participants during the interviews were explored (Lysack et al., 2017). Careful consideration was given to the format of questions in the interview schedule, allowing trust and rapport to be developed between the researcher and the participant, as these factors have been shown to be important in successful interviewing (Sivell et al., 2019). Pilot interviews have been demonstrated to be beneficial in informing interview guides in advance of formal data generation (Kim, 2010). Within this study, two pilot interviews were conducted with occupational therapy students with prior experience of conducting sessions using telehealth. These interviews were used to test and refine the interview guide questions based on participant feedback and the reflections of the researchers. Data generated from pilot interviews was not included in the analysis.

### Participants

The Health Service Executive [HSE], Ireland’s public health service, is divided into nine community healthcare organisations [CHOs] (HSE, n.d.), which deliver services to service users in their own communities. CHOs are further divided into smaller community healthcare services based on geographical location. Occupational therapists were recruited from a community mental health service within the HSE. Approximately twenty occupational therapists work as part of a multidisciplinary team within this service. These therapists are split between four practice sites across the geographical area covered by the service. Prior to the COVID-19 pandemic, these therapists typically delivered services within their respective healthcare centres, as well as in the communities of their clients. However, following the onset of the COVID-19 pandemic, a high proportion of occupational therapy sessions within this service were required to be delivered using via telehealth, commencing in approximately March 2020.

Purposive sampling was used within this service to target occupational therapists with experience of delivering mental health occupational therapy via telehealth. Purposive sampling aims to recruit participants with experiences which are relevant to the research questions being asked (Bradshaw et al., 2017). A flyer, information sheet, and consent form were circulated to all mental health occupational therapists within the mental health service by the occupational therapy manager. Occupational therapists who delivered a telehealth intervention within this service in the previous 12 months were invited to contact the researchers to express their interest in this study. Four occupational therapists were recruited for this study.

### Data Generation

Data was generated through one individual, semi-structured interview with each participant. Participant interviews were scheduled via email. Written, informed consent was obtained from each participant prior to their interview. Verbal consent was also obtained at the Cbeginning of each interview. Interviews were conducted remotely to comply with public health advice related to COVID-19. Participants had the choice of taking part in an interview via phone call or using a secure videoconferencing platform, depending on their needs and preferences. All four interviews were conducted using the Microsoft Teams videoconferencing platform. Online interview formats have been noted as comparable to in-person interviews for data generation and can increase flexibility for participants who may have difficulty taking part in studies due to time constraints or location (Gray et al., 2020; Janghorban et al., 2014).

Each interview was approximately 45 minutes in length and conducted by one of the two primary researchers. One researcher was present for each interview, as limiting power imbalances in the researcher-participant relationship is important during the interview phase to build trust and to ensure rich data is generated (Karnieli-Miller et al., 2009). Interviews were audio-recorded and transcribed verbatim. Both primary researchers contributed to the transcription of the interviews. Each transcript was checked for accuracy by both researchers. All identifying participant information was removed during the process of transcription. To support reflexivity, field notes were taken during all interviews (Shenton, 2004).

### Data Analysis

Thematic analysis (Braun & Clarke, 2006) was used to detect, analyse, and report patterns within the data generated. This method of analysis was used as it enables the organisation and description of the data in rich detail (Braun & Clarke, 2006). Familiarisation with the data was achieved through transcription of the interviews by the researchers, which is recommended to assist with the analysis of qualitative data (Braun & Clarke, 2006).

In the first stage of analysis, the participating occupational therapists’ transcripts were read and re-read to gain a full understanding of the data and to ensure familiarity with the content. Initial thoughts and ideas for coding were noted and were returned to in subsequent steps. During the second stage, preliminary codes were produced. Each transcript was coded independently by each of the researchers. Discussions involving reflection were held between researchers to reach consensus. During the third stage, codes were sorted into potential themes. During the fourth stage of analysis, these themes were reviewed by both researchers and the research supervisor to ensure they reflected the content of the transcripts of the occupational therapists as a whole. Stage five involved defining each theme and attributing each a title for use within the findings.

### Rigour and Trustworthiness

Rigour in qualitative research ensures that experiences and meaning are accurately captured within the data generated (Lysack et al., 2017). To ensure rigour and trustworthiness within qualitative research, consideration must be given to the credibility, transferability, dependability, and confirmability of the study (Carpenter & Suto, 2008; Nassaji, 2020). Several strategies were used within this study to ensure rigour and trustworthiness. Independent coding of the transcripts by each researcher supported investigator triangulation and compensated for single-researcher bias, thus supporting the credibility of the findings (Curtin & Fossey, 2007; Korstjens & Moser, 2018). Prior to establishing the finalised findings, a process of confirmation was conducted with the research supervisor to further increase the credibility and confirmability of the study’s findings (Shenton, 2004). An audit trail was used to ensure the confirmability and dependability of findings (Korstjens & Moser, 2018). A reflective journal was kept throughout the course of the study to ensure researcher reflexivity and to address potential biases of the researchers affecting the findings (Curtin & Fossey, 2007).

### Ethical Considerations

Ethical approval for this study was granted by the Social Research Ethics Committee, University College Cork, in January 2021. To ensure participant confidentiality, all initial contact with participants was made through an identified gatekeeper to limit the amount of personal and demographic information accessible to the researchers. All participants were provided with a consent form and an information sheet describing the purpose of the study as well as participation requirements. Participants were informed in advance of their interview, as well as during the data generation phase, that they could withdraw from the study at any time, up to two weeks after their interview had been conducted. All data was stored on an encrypted laptop, and all audio recordings were destroyed following interview transcription, to further ensure participant confidentiality.

## Findings

During the iterative process of data analysis, three core themes emerged relating to the experiences of occupational therapists in transitioning to telehealth during the COVID-19 pandemic, as displayed in Table I. Identifier codes have been applied to all quotations to ensure confidentiality. Quotes from the four participants will be accompanied by the identifier codes ‘OT1’ to ‘OT4’.

**Table I.**
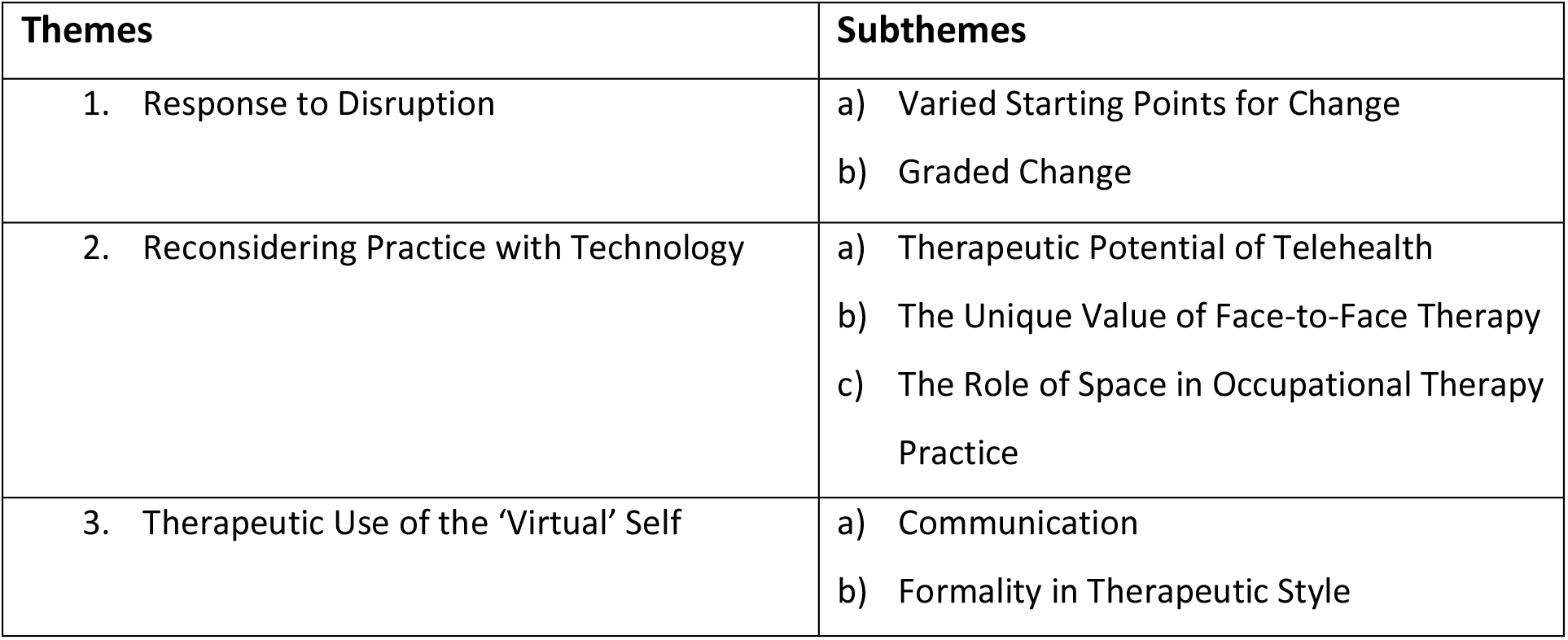
Themes and Subthemes.

### Response to Disruption

The transition to telehealth was described by participants to be a disruptive event, as a reactive change occurred across the service in response to the COVID-19 pandemic. Occupational therapists reported having limited exposure to telehealth as a model of service delivery prior to COVID-19, with insubstantial technological infrastructure in place to facilitate this change. Therapy services were reported to have been disrupted, and a process of graded change in the initiation of telehealth implementation was described. The two subthemes associated with this service’s response to disruption were varied starting points for change and the process of graded change.

#### Varied Starting Points for Change

It emerged from the data that each telehealth user commenced the transition to telehealth with a unique foundational level of preparedness to engage in telehealth services. Participants described how each individual occupational therapist and service user had their own starting point in transitioning to telehealth, based on a number of personal factors, which in turn influenced their level of success and the time taken to engage in telehealth services. Participating occupational therapists reported that factors such as the user’s level of apprehension, comfort with technology and technological skills influenced their ability to engage with telehealth. All participants outlined how the user’s level of familiarity and previous experience with technology impacted upon their ability to engage in telehealth sessions.

> *I was comfortable using it, you know. I use it regularly… things were going wrong… and the person on the other end wasn’t familiar with… online platforms, I was able to problem solve quickly…* (OT3)

The starting point for each occupational therapist or service user appeared to determine the steps needed to support their transition to telehealth, and also indicated the ease with which the user would engage with online services.

> *…if they’re somewhat IT literate, that is a help* (OT1)

#### Graded Change

Participants described the process of making the transition to telehealth as a process of graded, incremental change. Incremental changes were made towards telehealth engagement, with the initial steps taken based on the user’s starting point. Therapists’ understanding of activity analysis appeared to support clients’ transition to telehealth, as well as their own transition, through the identification of facilitators and barriers.

> *As OTs, one of our skills is finding the barrier to occupation… Using technology, that is kind of occupation… We have that skill as OTs to assess*… *the person, the environment, the occupation, kind of see what where the breakdown is and see what we can do to… help the person do it*. (OT4)

Increased experience gained by participants and their clients, through gradual exposure to technology platforms and progressive building of their skills, facilitated successful telehealth engagement. Therapists interviewed as part of this study outlined how client engagement was purposely graded, beginning with aspects of telehealth which were deemed to be less complex, such as phone calls, and progressively built towards elements which were more complex, such as videoconferencing sessions. Occupational therapists described strategies used to develop the skills required for telehealth for both themselves and their clients, such as education, use of phone calls as an initial step to remote interaction with the service, and short trials of videoconferencing platforms. These strategies were reported to have been beneficial in supporting the engagement of both occupational therapists and service users.

> *I would offer to, you know, for… the first couple of minutes of a session just to… try it, come up on the screen, and then we can end it and just finish… the session on a* [phone] *call*… *Those sort of things seemed to help people because they, they could do it in a safe way. They knew there wasn’t any pressure to stay on the video call…* (OT3)

Occupational therapy service provision was also incrementally transitioned to online platforms, beginning with practice elements of least complexity, such as check-in sessions with individual clients, and gradually incorporated elements of increasing complexity, such as online groups. All occupational therapists interviewed used individual sessions to increase their own comfort and familiarity with telehealth, as well as that of their clients. The transition of occupational therapy groups online occurred later in the process of change, when occupational therapists had gained experience with individual online sessions.

> *People are kind of just feeling*… *a little bit more confident from having done… the individual sessions that*… *I think we can actually do this now, you know, in a group session*. (OT2)

### Reconsidering Practice with Technology

Through a reactive transition to telehealth, therapists were given the opportunity to evaluate the role of technology within the service, and where this technology may be integrated into a face-to-face model of service delivery in the future. The therapeutic potential of telehealth, the unique value of face-to-face therapy, and the role of space in occupational therapy practice presented as subthemes relating this theme.

#### Therapeutic Potential of Telehealth

All participant interviews highlighted that there is inherent value in the use of telehealth within community mental health occupational therapy services. Benefits reported by the participating occupational therapists can be categorised as increases in flexibility, accessibility, and the efficiency of services when delivered via telehealth. Therapists described how the delivery of sessions remotely reduced demands placed on clients to attend the session, with decreased incidences of non-attendance following telehealth implementation. All participants reported that telehealth sessions offered better alignment with clients’ roles and routines, with an increased feasibility of shorter sessions to accommodate for other commitments.

> *Because our hours are kind of eight til four, nine to five… people who have work in those hours might end up missing sessions because of that. But being able to do something online means they can link in on their lunch break or they can link in… before they start work…* (OT3)

Participating therapists acknowledged that telehealth has the potential to increase the efficiency of a service, with reduced travel time required for service users as well as professionals. Telehealth facilitated increased collaboration across the multidisciplinary team, as an increased number of meetings conducted using videoconferencing platforms could be facilitated, with reduced travel required to liaise with professionals based in different sites. Due to the recognition of this potential, all participants supported the idea that telehealth is *‘here to stay’* (OT1) and is likely to remain as a permanent component of practice in community mental health services going forward.

#### The Unique Value of Face-to-Face Therapy

Participants highlighted that there were specific components of practice which were of most value when conducted in-person. Observations and functional assessments are components of practice which therapists reported to be most precise when conducted in-person. Additionally, participating therapists reported that clients experiencing high levels of distress would benefit most from in-person sessions and may find telehealth sessions more challenging.

> *I don’t think it would work well with client groups who are extremely unwell. I think you have to have a level of wellness to be able to engage with the person on the screen* (OT1)

The therapists interviewed as part of this study suggested that the demands of attending a clinic session may present positive challenges for clients in some circumstances. The therapists reported that clinic attendance encouraged clients to engage in activities of daily living (ADLs), as well as occupations such as the use of public transport. Participating occupational therapists suggested that engagement in these occupations may support the wellbeing of some clients by contributing to the structure and routine of their schedules. All participants supported the idea that telehealth *‘won’t completely replace face-to-face’* (OT1), given the unique benefits presented by in-person therapy.

#### The Role of Space in Occupational Therapy Practice

The significance of the role of space within occupational therapy practice was re-evaluated through the use of technology, as occupational therapists reported engaging in therapy sessions from different locations, such as their homes. All participants outlined how the space in which a session occurs can add value to the therapeutic process, while some spaces may present challenges. It was widely reported by the participating occupational therapists that the clinic location adds a component of neutrality to the sessions and supports client boundaries and privacy. Therapists reported that, within this neutral space, the impact of clients’ roles and responsibilities, such as caring demands, were minimised.

> *it was hard for her… to kind of… make the time for the video call, to not be distracted by all the younger children in the house… the neutrality of being, you know, up here and removed from other kind of, you know, stressors or busyness in her life, you know, that, that would have been better for her*. (OT2)

Similarly, clients’ engagement with the occupational therapists in locations other than the clinic, using telehealth, reportedly added to the value of sessions in a unique way.

Occupational therapists interviewed as part of this study reported that some of their clients experienced increased levels of comfort in their home environment, which facilitated client engagement in the therapeutic process. Occupational therapists reported that the use of video calls allowed them to see the client’s home environment, which provided additional information to support and inform interventions.

> *I was actually very specifically working on ADLs with him and kitchen skills, so the brother was able to show me, they actually, so they actually brought the computer around the kitchen… You walk through the kitchen with them and you’re seeing what the difficulties are… where the goals are going forward…* (OT1)

### Therapeutic Use of the ‘Virtual Self’

The participants described a new understanding of the therapeutic use of self as it applies in a virtual context. Participants described how the demands of the virtual environment led to changes in communication, which in turn impacted upon the development of rapport. All therapists interviewed as part of this study reported a tendency to deliver a more formal and structured style of therapy while working with clients online.

#### Communication

Therapists included in this study reported altered styles of communication occurring online as a result of ‘*difficulties with connection*’ (OT3) as well as a reduction in visual cues. Participating occupational therapists reported that poor internet connection and call lagging led to a ‘*disjointedness*’ (OT2) in conversation. All participants reported that the reduction in visual cues and limited visibility of body language also limited the therapists in gaining a full understanding of the experiences of their clients.

> *Body language, getting a sense from the person, you know…they could have other kind of anxious body language pieces going on that you can’t see or aren’t visible on the phone or the video call* (OT3)

The difficulties experienced in communicating led to the limiting of conversation to the most essential messages, with a significant reduction in informal interactions, or ‘*small talk’* (OT3), as reported by participating therapists. Therapists included in this study described the manifestation of these communication difficulties as a ‘*disconnect*’ (OT2) in rapport with their clients.

Despite the reported communication difficulties, participating therapists reported that a conscious inclusion of ‘*informal time*’ (OT3) within telehealth sessions had the potential to facilitate the development of therapeutic rapport. This technique was a strategy intentionally applied to counteract the changes in communication which were reported to occur within telehealth sessions.

#### Formality in Therapeutic Style

Therapists interviewed as part of this study described using a style of therapy in telehealth sessions which was more structured and *‘formalised’* (OT3) in nature than in face-to-face sessions. The participating therapists reported that the reduction of informal interactions resulted in more structured and goal-oriented sessions.

> *it feels effective and it feels like we need to achieve something here and we’re both here for a reason… it feels formal in a way – probably in a good way*. (OT1)

Participating therapists described how online sessions were suited to working on *‘concrete, specific goals’* (OT2), rather than providing *‘generic support’* (OT2). Assessments and interventions which had a specific structure, such as the Occupational Circumstances Assessment Interview and Rating Scale (Version 4) (Forsyth et al., 2005), and psychoeducational interventions were reported to transfer well to online formats, with therapists describing an increased level of preparation and planning required for online sessions.

> *I would have been more prepared, kind of, with my notebook, specifically what we were going to look at*. (OT2)

## Discussion

The findings of this study provide a new understanding of the process of change which occurs as an organisation transitions to telehealth in response to the disruption of services. This study aimed to describe the process of transition to telehealth as it occurred within a community mental health setting, as well as to explore the opportunities and challenges presented by the implementation of a remote model of service delivery. This study further aimed to examine the transfer of the occupational therapists’ skills to telehealth sessions. This study presents novel findings relating to telehealth in mental health occupational therapy practice. While the prevalence of research relating to telehealth in occupational therapy has been increasing over the past decade, there has been a limited focus on telehealth in mental health occupational therapy to-date.

Disruption within healthcare occurs when an organisation must change the delivery of its services in response to external disturbance (Kaufman & Grube, 2015). The delivery of services within the participating organisation was disrupted by the onset of the COVID-19 pandemic. The organisation was required to incorporate a telehealth model of service delivery to enable the continued provision of services in alignment with public health guidelines. Kaufman and Grube (2015) signify the need for a *‘blueprint’* (p.54) for change when incorporating new models of service delivery in response to disruption. As illustrated in the findings, without such a blueprint, therapists were required to gain the necessary knowledge pertaining to online service delivery through experiential learning with continued engagement in telehealth. Therapists developed this knowledge of telehealth by first delivering the least complex aspects of practice, such as check-in phone calls, and then progressing to the delivery of sessions of increased complexity, such as online group interventions. The knowledge gained through the delivery of less complex sessions informed the planning and implementation of more complex elements of practice. As the use of telehealth was limited in Ireland prior to the pandemic (Lolich et al., 2019), a framework outlining the steps necessary to transition a service online was not available to this organisation. Occupational therapy services may benefit from the development of a framework guiding the transition to telehealth.

The findings of this study suggest that the process of change is evident in how occupational therapists integrate telehealth solutions into their practice. The commitment to a *‘new view of health care’* (p. 53) may facilitate a successful response to disruption (Kaufman & Grube, 2015). However, resistance to telehealth implementation has been reported within the existing literature and has been suggested to be a barrier to change (Lolich et al., 2019; Morris et al., 2019). Resistance was evident in the findings of this study through the apprehensions participants reported to have experienced in advance of telehealth implementation. Participants’ lack of familiarity, experience, and knowledge relating to telehealth in advance of its’ implementation amplified apprehensions, and increased resistance to remote service delivery. However, theme 1 within the findings, detailing the service’s response to disruption, illustrates an increased acceptance of telehealth following a period of online service delivery. In the context of this study, although the transition to using technology was not anticipated, therapists came to recognise how practice could be enhanced using technology, such as providing increased accessibility and flexibility for service users. Their commitment to this new, remote model of service delivery was demonstrated in the findings through the participants’ desire to maintain remote services as a permanent component of practice in the future.

The findings of this study suggest that occupational therapists would favour a blended approach to practice going forward, with components of both face-to-face therapy and telehealth integrated into one service delivery model, due to the efficiency and alignment with clients’ roles and routines which telehealth can provide. Literature within occupational therapy relating to telehealth recommends a ‘*hybrid model*’ (p.38) including face-to-face therapy in combination with telehealth (WFOT, 2014). However, references to such blended models of practice within occupational therapy are somewhat ambiguous in defining the exact formulation of these service delivery models, without clear guidance on how these two modalities could be merged within a specific healthcare service. The findings of this study illustrate how occupational therapists identified aspects of practice which were augmented through the use of telehealth, such as increased efficiency in conducting check-in sessions, with reduced time commitments for clients, and effective delivery of psychoeducational sessions. Theme 2 of the findings also displays how, through use of technology, occupational therapists could identify unique benefits of face-to-face sessions, such as conducting functional assessments and working with clients experiencing high levels of distress. This outlines how occupational therapists might begin to create a more specific model for telehealth services going forward, with a view to facilitating service user choice, increasing efficiency and improving client engagement. As indicated from the literature review, the creation of such an approach to practice would increase the accessibility of services (Hines et al., 2019; Kuravackel et al., 2018; Little et al., 2018; Rosenbek Minet et al., 2015; Scriven et al., 2019). Identification of aspects of the occupational therapy process which are best suited to a particular mode of service delivery could contribute to the development of a framework for a blended model of service delivery in community mental health settings. The creation of such a framework, outlining the place of both face-to-face and online modalities, could have the potential to guide the restructuring of existing service delivery models to incorporate a more blended approach to practice in the future.

Findings from this study suggest the therapeutic “use of self” changes as practice transitioned to a remote model of service delivery. The therapeutic use of self is the process in which occupational therapists develop and maintain therapeutic relationships with service users (AOTA, 2020). Distinct skills reported to contribute to the development of this relationship include effective communication and interpersonal skills (McKenna, 2017). However, existing literature relating to the therapeutic use of self is based on face-to-face models of service delivery and this concept has yet to be explored in a virtual context. As illustrated in the findings of this study, technology related difficulties such as poor connectivity prevented therapists’ use of particular communication skills, such as informal conversation and the use of non-verbal cues, thus imposing an increased formality of communication between therapist and client. As reported by the participating therapists, the reduction in the use of these communication skills and increased formality presented challenges to the development of the therapeutic relationship. Considering that the current technology available to occupational therapists does not support the application of their interpersonal skills, therapists must consider what alternative skills can be used to foster trust and develop rapport with clients in virtual settings. In this study it was evident that less complex elements of practice, such as check-in sessions, transitioned to telehealth platforms with relative ease, whereas more complex practice elements, such as group intervention, took longer and required more experience (see Theme 1 in Findings). Therefore, given its complexity, it remains to be seen if all skills pertaining to the therapeutic use of self, such as informal conversation and non-verbal communication may be transferred fully to telehealth platforms as therapists gain further experience and knowledge relating to remote service delivery. Further research is required to delineate the skills involved in the development of therapeutic relationships in remote settings and to explore the therapeutic use of self as it occurs in a virtual context.

## Limitations

The small sample size recruited for participation in this study may limit the generalisability of the findings. Further research is required with a larger number of participants, with particular emphasis on representing and exploring the service user experience. In the interest of maintaining confidentiality and anonymity, demographic information was not gathered from the participants of this study. However, this may have had an impact on the transferability of the findings. The inclusion of demographic data in future studies may be beneficial in improving the trustworthiness of available evidence in this area. Interviews were conducted remotely due to COVID-19 restrictions. The remote methods used to interview participants may have skewed the representation of participants within the study to those who were already proficient in technology use. Further studies using in-person research methods may accommodate the inclusion of individuals with lower levels of technology proficiency, in order to provide a more comprehensive description of the experience of transitioning to telehealth within community mental health settings. This study was conducted while the participating service was going through a process of transition, and views of participants may change as they gain further experience with telehealth. Future studies conducted with services who have had more prolonged experience with telehealth may be beneficial in further illuminating this process of transition within community mental health practice.

## Implications for Practice

The COVID-19 pandemic offered occupational therapists in Ireland a unique opportunity to identify the potential for telehealth within their practice, which is likely to become a permanent feature of practice going forward. As unrestricted delivery of in-person occupational therapy returns, therapists must consider the elements of practice which are best suited to remote delivery, using the experience and knowledge gained through telehealth engagement during the pandemic, to inform blended service delivery models going forward. Due to the lack of an available framework to guide the transition to telehealth, gradual, incremental changes should be implemented by therapists in transferring their practice to remote service delivery, starting with the least complex elements of their practice, and gradually incorporating the elements of most complexity. Therapists must assess their knowledge and skills relating to telehealth, as well as those of their clients, in advance of telehealth implementation, to determine the steps required to facilitate this transition. The development of a framework guiding this change, informed by the collective experience gained by therapists during the pandemic, could support services in making this transition in the future. Additionally, further exploration of the service user experience is imperative to inform future service delivery. Further research should also be conducted with larger sample sizes and across multiple services, to increase the generalisability of findings across occupational therapy practice.

## Conclusion

The COVID-19 pandemic has led many occupational therapists in Ireland to re-evaluate the role which technology may play in their practice. The use of telehealth in mental health occupational therapy has been shown to present unique opportunities for practice, with increased flexibility for clients and heightened efficiency for therapists. Telehealth is likely to remain a permanent component of practice going forward, as a blended approach to service delivery is adopted, with increased frequency of remote sessions. The process of graded change described by the participants of this study suggests that a gradual transition would be required to incorporate telehealth into existing occupational therapy services. Due to increased telehealth uptake during the COVID-19 pandemic, occupational therapists have gained the experience and knowledge of telehealth necessary to identify its inherent potential, which will inform the development of blended service delivery models in the future.

Furthermore, the formality of the therapeutic relationship warranted by remote platforms challenges occupational therapists to adapt their skills to foster trust and rapport with clients. This is particularly challenging in a remote context which is not supportive of communication techniques which typically assist in the development of this relationship in a face-to-face context, such as the use of non-verbal communication. Therapists must consider what skills can be used to facilitate the development of a therapeutic relationship in a remote context. These skills may then be used to conceptualise the therapeutic use of the ‘virtual’ self.

## Data Availability

Data will be made available on request by the corresponding author.

